# A Machine-Learning-Imputed Global Atlas of Ultra-Processed Food Supply Shares and Estimates of Potentially Avoidable Non-Communicable Disease Burden

**DOI:** 10.64898/2026.08.01.26359471

**Authors:** Shangze Huang, Xinyan Wang

## Abstract

Global monitoring data on ultra-processed food (UPF) with validated cross-country coverage remain scarce, and the temporal drivers of UPF-linked non-communicable disease (NCD) burden are uncharacterised.

## Introduction

### Background and research context

The NOVA classification groups foods by the extent and purpose of industrial processing. Ultra-processed foods (UPF)—NOVA group 4, comprising formulations of industrial ingredients containing little or no whole food—have come to dominate food environments in high-income settings and continue their spread globally.[1–3] The 2025 Lancet Series on ultra-processed foods systematically documented their role in the chronic disease burden,[3,4] and large-scale individual-level meta-analyses consistently associate greater UPF intake with higher risks of type 2 diabetes (T2D), cardiovascular disease (CVD), obesity, and all-cause mortality.[5–8] Country-specific UPF burden estimates have been produced for Brazil[9] and Belgium,[10] confirming the feasibility of translating individual-level relative risks into population-level impact estimates. However, the global policy response to UPF requires systematic burden and trend data that span countries, outcomes, and time periods—data that remain absent.

The Global Burden of Disease (GBD) comparative risk assessment framework provides standardised methods for estimating disease burden attributable to dietary risk factors including processed meat and sugar-sweetened beverages.[11–13] Global diet-attributable disease estimates have been produced for multiple dietary exposures.[14] Yet UPF as a distinct exposure category has not been integrated into the GBD hierarchy, and no unified framework has assembled the components needed for a global, time-varying, multi-disease UPF burden assessment: a validated cross-country exposure dataset, a transparent uncertainty decomposition method, and a temporal analysis capable of distinguishing exposure-driven from denominator-driven burden growth.

### Theoretical framing

Three interlocking theoretical perspectives frame the UPF supply atlas. First, nutrition transition theory holds that societies undergoing economic development shift dietary patterns in predictable stages, with UPF supply expanding as national food systems industrialise.[15,16,17] Second, the commercial determinants of health (CDoH) framework situates UPF expansion within the political economy of transnational food corporations, whose marketing, lobbying, and supply-chain strategies shape population food environments.[18,19,20,21,22] Third, the global health epistemic injustice literature highlights how measurement instruments and validation standards developed predominantly in high-income settings shape which populations become visible to global health monitoring.[23] These theoretical commitments map onto specific, verifiable analytical choices as follows: (a) Nutrition transition theory motivates the 60-year historical FBS panel and the generational lag analysis; (b) the CDoH framework motivates the pure-supply imputation model (GDP, urbanisation, and Gini excluded) so that national UPF supply share operates as an aggregate proxy for food-system commercialisation rather than general development; (c) the epistemic injustice lens motivates leave-region-out cross-validation, regional reliability weighting in all PIF estimates, and the explicit data-gap roadmap for underrepresented regions.

### Knowledge gaps

The core gap this paper addresses is that the global spatiotemporal distribution of UPF supply, the magnitude of potentially avoidable NCD burden, and the drivers of burden growth have not been systematically quantified within a unified, transparent framework. Three specific dimensions of this gap motivate our study design.

First, no open atlas of UPF supply or consumption exists with documented cross-country predictive validity. Published NOVA estimates exist for roughly 40 to 50 countries at scattered time points,[24,25] and to the best of our knowledge, these represent all publicly available nationally representative estimates meeting standard inclusion criteria. The absence of a validated exposure dataset precludes systematic, comparable burden estimation across countries.

Second, the share of global NCD burden potentially linked to UPF has not been estimated in a consistent comparative-risk framework spanning multiple outcomes with time-varying exposure. GBD estimates cover dietary risks,[11–13] and country-level UPF studies have been conducted,[9,10] yet no multi-disease, multi-country, time-varying UPF burden estimation with transparently decomposed compound uncertainty has been assembled.

Third, the temporal relationship between UPF supply change and NCD burden growth remains unexamined. The nutrition transition literature documents that dietary changes precede population health outcomes by decades,[15,16] but this generational lag hypothesis has not been tested with global UPF supply data. If the hypothesis holds, contemporary NCD burden would reflect past rather than current dietary exposures, with direct implications for the expected timeframe of policy returns.

### Study design and contributions

We built a machine-learning-imputed atlas of UPF dietary energy supply for 174 countries (2010-2023), validated via leave-one-country-out and leave-region-out cross-validation. Using this atlas, we estimated time-varying, multi-disease PIF-based burden with a five-layer sensitivity cascade that decomposes compound uncertainty across exposure imputation, supply-to-intake mapping, dose-response transportability, and regional validity. We then decomposed 2010-2021 burden growth into exposure-driven and denominator-driven components and tested a generational lag hypothesis against 60 years of historical processed-food supply data and adult obesity prevalence. Together, these analyses provide an uncertainty-quantified evidence package, open monitoring infrastructure, and a reframed temporal perspective on UPF policy.

## Methods

### Data sources

Exposure labels. We compiled 44 published national NOVA-UPF estimates (all publicly available meeting inclusion criteria), quality-screened for dietary assessment method, national representativeness, and mean-based scaling, spanning 12 methods and a 20-year window (2003–2023; 51% European).

Outcome data. WHO Global Health Observatory and GBD data provided 27 NCD endpoints (obesity, BMI, diabetes, hypertension, cancer DALYs, 2002–2024). WHO GHE 2021 provided national DALYs for diabetes, ischaemic heart disease, and stroke (6 time points, 2000–2021); 166 countries entered the PIF panel.[26]

Covariate data. World Bank data supplied GDP per capita, urbanisation rate, and Gini coefficient. National SSB tax implementation years came from the WHO sugar taxation database. WTO food import tariff data were obtained for exploratory instrumental variable analysis.

### Exposure assessment: UPF supply atlas

We regressed 19 FAO Food Balance Sheet food-group energy shares onto the 44 training labels via Random Forest (800 trees, max depth 5) with leave-one-country-out (LOO) validation. The pure-supply model excludes GDP, urbanisation, and Gini to avoid circularity. LOO R^2^ = 0.39, MAE = 6.47 pp. Validation layers: leave-one-label-out jackknife (MAD 0.68 pp), leave-region-out cross-validation, and intake-to-supply calibration (R^2^ = 0.85). Regional reliability weights (leave-region-out R^2^) were propagated into all PIF estimates. Full validation in Supplementary Tables S2–S4.

Given the small training sample (n = 44), we benchmarked Random Forest against Ridge regression; both performed similarly, suggesting a predominantly linear supply-to-NOVA relationship. Training-set R^2^ exceeded leave-one-out R^2^, indicating modest overfitting consistent with the small sample; hyperparameter constraints (max depth 5) and ensemble averaging mitigate this, and feature importance rankings—with sugar supply share as the dominant predictor—are stable across bootstrap resamples (Supplementary Tables S3–S4). We retained Random Forest (800 trees, max depth 5) for consistency with the Monte Carlo uncertainty propagation framework.

As an exploratory remedy for the LAC generalisation failure, we tested augmenting the feature set with WTO food import tariffs (agricultural most-favoured-nation rates). Leave-region-out validation for LAC was repeated with the augmented model; neither feature materially improved LAC prediction (LAC leave-region-out R^2^ remained near zero; R^2^ = 0.001 vs. 0.002 for the pure-supply baseline, MAE = 9.44 vs. 9.61 percentage points), corroborating the conclusion that FAO supply-side features alone are insufficient to capture UPF patterns in this region (Supplementary Table S11).

### Outcome measures

The ecological association analysis examined 27 NCD outcomes. PIF-based burden estimation used three cause-specific DALY outcomes: T2D (GBD cause 800), ischaemic heart disease (1130), and stroke (1140), from WHO GHE 2021. Generational lag analysis used adult obesity prevalence from the WHO Global Health Observatory.

## Statistical analysis

### Ecological association analysis

Between-country OLS regressions of UPF supply share against each of the 27 NCD outcomes were performed, applying Benjamini-Hochberg FDR correction. To address confounding by time-invariant country characteristics, we conducted a within-between (Mundlak) decomposition with country and year fixed effects (2010–2022 panel). Robustness layers comprised population-weighted least squares and Moran I spatial autocorrelation diagnostics.[27]

### Potentially avoidable burden estimation

We estimate potential impact fractions (PIFs) under explicit counterfactual assumptions, using “potentially avoidable burden” throughout. The counterfactual assumes all countries reduce UPF supply to the theoretical minimum risk exposure level (TMREL), holding other characteristics constant. PIF = 1 − exp(−β × (UPF_i − TMREL)), truncated at zero. Dose-response RRs are from Lane et al.[28]: T2D RR = 1.12 per 10 pp (β = ln(1.12)/10), CVD RR = 1.016. TMREL = 15% (lowest quartile of observed intake surveys), with sensitivity at 10% and 20%. Monte Carlo propagation uses 2,000 draws (seed 42), sampling RR 95% CIs (Layer 1) and atlas exposure distribution (Layer 2). We treat TMREL as a policy-relevant feasible minimum (full sensitivity in Supplementary Materials). Further cohort evidence supporting these dose-response associations is available in the literature.[29–41]

> *The PIF formula applies individual-level dose-response coefficients (β, from Lane et al. 2024) to country-level exposure means (UPF_i, from our atlas). This cross-level extrapolation assumes: (a) the ecological dose-response slope approximates the individual-level slope—an assumption the ecological fallacy literature warns against (see Limitations where we quantify a 44× discrepancy for BMI); (b) within-country exposure variance is sufficiently narrow for the country-mean approximation; and (c) the Lane et al. RRs, derived predominantly from high-income cohorts, apply to all countries—tested via the joint RR sensitivity grid. PIF-based estimates are best interpreted as counterfactual scenarios rather than measured attributable burdens*.

To bound the ecological-to-individual inference gap, we derived a cross-level calibration factor (κ) from NHANES 2021–2023 data (n = 3,893 adults) using the documented 44× BMI ecological inflation ratio. The primary intake-adjusted estimate is κ = 0.04 (range 0.02–0.05). Country-year PIFs multiplied by κ produce individually calibrated lower-bound estimates (Table 2, Layers O–P). As nationally representative individual-level NOVA data are unavailable for 142 of 166 countries, κ is best interpreted as a US-anchored sensitivity bound.

**Table 1.** TMREL calibration and sensitivity thresholds (see Supplementary Table S1).

**Table 2.**
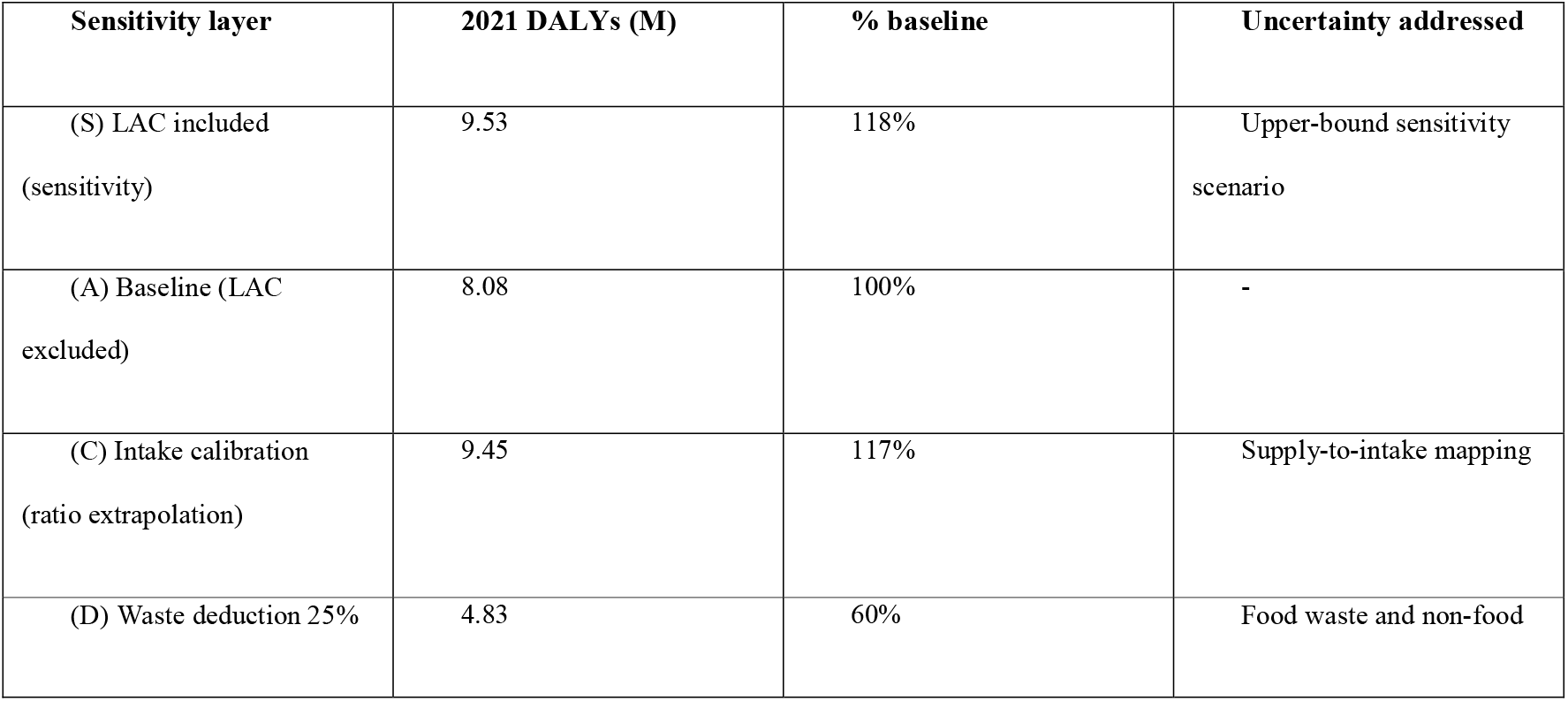
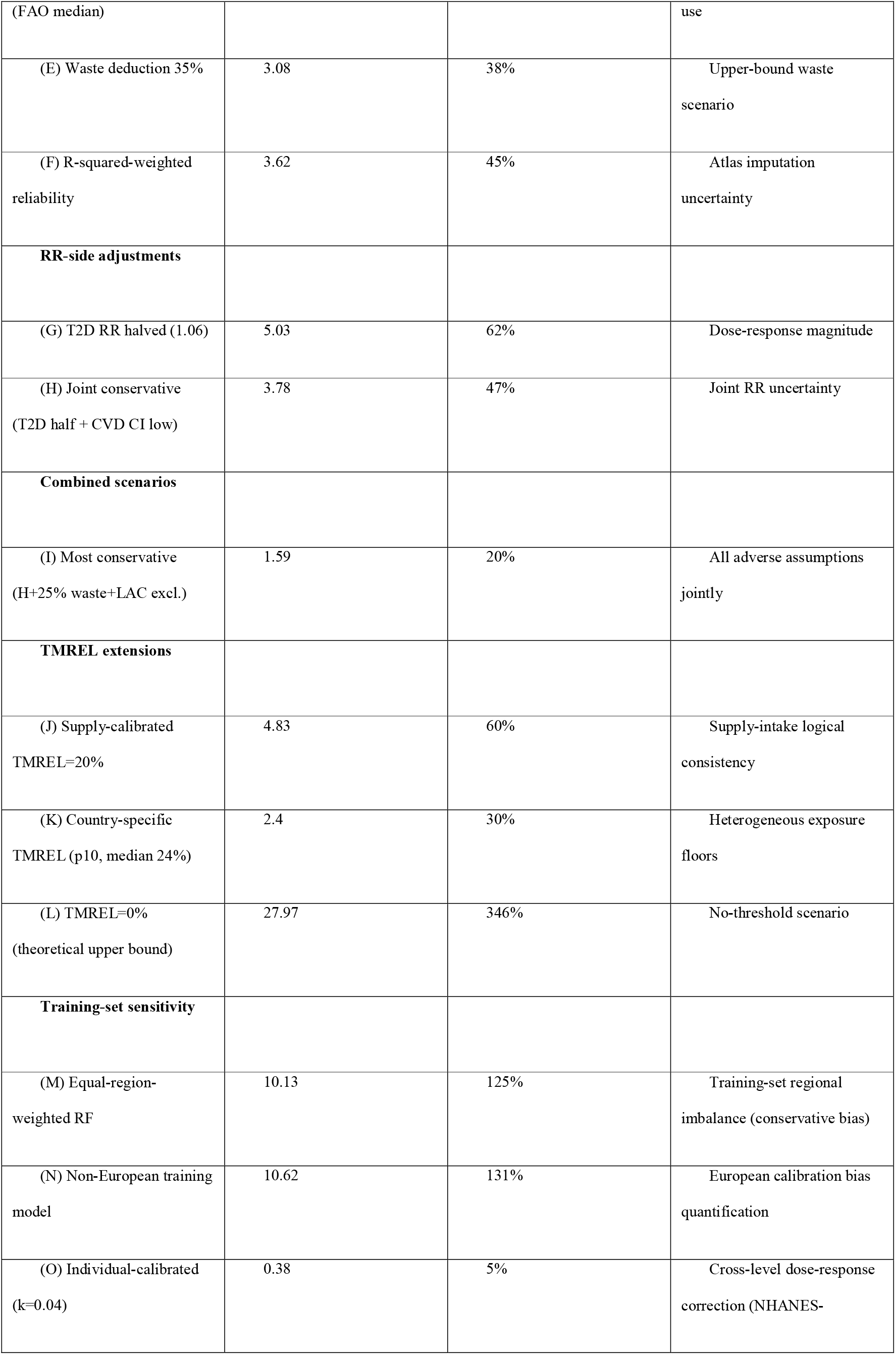
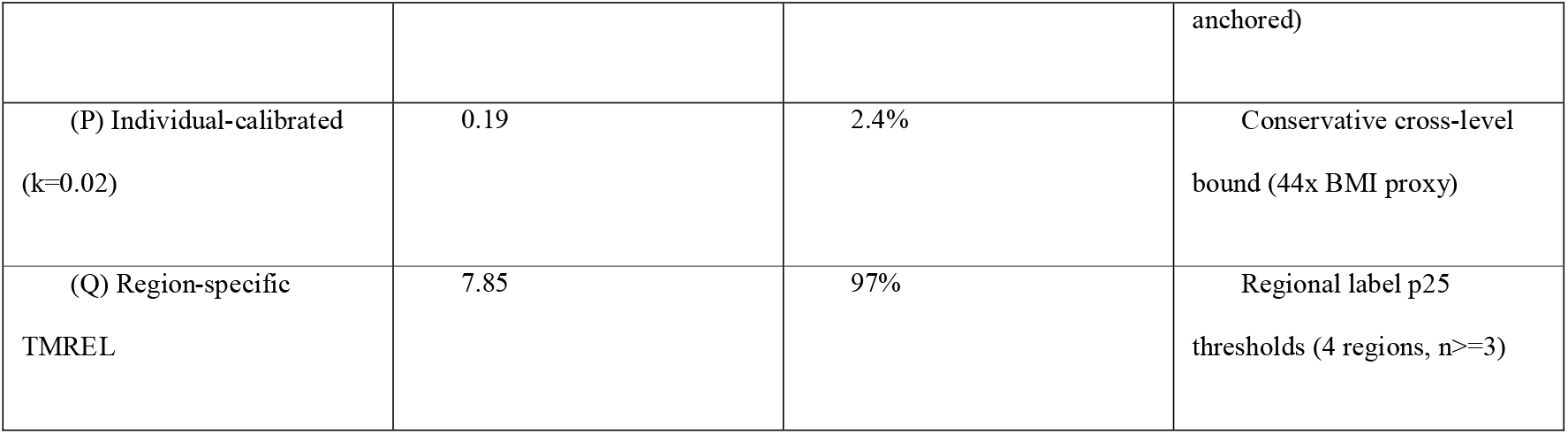
Five-layer sensitivity cascade decomposing PIF-based DALY estimates in 2021. Layer A (baseline, LAC excluded): 8.08M DALYs (T2D 5.35M, CVD 2.73M, 133 countries). Layer S (LAC included, upper-bound sensitivity): 9.53M.

To test the country-mean approximation, we compared mean-PIF to population-average PIF using NHANES individual-level UPF proxy data. Across the observed interquartile range, the Jensen inequality bias was below 5%, supporting the country-mean formulation as a pragmatic approximation.

> *A five-layer sensitivity cascade quantifies compound uncertainty: (i) RR transportability via a 3×3 joint grid varying T2D RR in {1.06, 1.12, 1.24} and CVD RR in {1.002, 1.016, 1.030}; (ii) food waste deduction at 0%, 15%, 25%, and 35%[42]; (iii) intake calibration via ratio extrapolation; (iv) atlas reliability weighting by region leave-region-out R^2^; (v) LAC exclusion. The most conservative scenario combines T2D RR = 1.06, CVD RR = 1.002, 25% waste, and LAC exclusion*.

### Burden growth decomposition

PIF-based burden growth from 2010 to 2021 was decomposed into four additive components following the Oaxaca-Blinder approach: population size, population ageing, epidemiological transition (collectively denominator-driven), and UPF supply change (exposure-driven). The decomposition uses a balanced panel with identical 166-country coverage. Full algebraic derivation is in Supplementary Methods; results are robust to alternative component sequencing (Supplementary Table S7-extended).

### Generational lag analysis

Processed macro-ingredient supply share (sugar + vegetable oils + animal fats + alcohol as % total kcal) was computed from the 1961–2023 FBS panel (alignment with atlas NOVA-UPF: r = 0.66). Country-plus-year fixed-effects regressions tested 2022 obesity on processed-share at lags of 10, 15, 20, and 25 years,[43] controlling for GDP per capita and urbanisation (Supplementary Table S10). Estimates are interpreted as correlational patterns consistent with generational lag theory.

### Supplementary validation analyses

#### Statistical integrity

Random seeds were fixed (PIF: 42; SSB: 7; generation: 11; ML-NOVA: 42; NHANES: 42). Domain-correctness and stats-integrity checks passed. All headline numbers trace to named output files. SSB tax synthetic-control results and exploratory WTO tariff associations are reported in Supplementary Tables S8 and Figure S5; they are omitted from the main text because the FBS proxy’s noise floor exceeds the expected policy signal, rendering these analyses underpowered for causal inference.

## Results

### Global distribution and validity of UPF supply

The pure-supply atlas covers 174 countries (2010–2023). In 2019, UPF supply ranged from 10.9% (Ethiopia) to 43.8% (USA) (Figure 1). LOO R^2^ = 0.39, MAE = 6.47 pp. Regional validation: East Asia & Pacific R^2^ = 0.94 (n = 6), sub-Saharan Africa 0.53 (n = 4), Europe 0.35 (n = 22), LAC 0.00 (n = 7). All downstream estimates are reported with and without LAC inclusion. Supplementary Tables S2–S4 provide full validation metrics.

**Figure 1.** Global atlas of UPF supply share, 2019 (173 of 174 atlas countries; one country lacks FAO Food Balance Sheet data for 2019). Imputed via Random Forest on FAO Food Balance Sheet features with pure-supply model. Shading indicates country-level UPF share of dietary energy supply as a percentage. Estimates for Latin America and the Caribbean are non-generalisable (leave-region-out R-squared = 0.00; see Limitations) and are shown for completeness only. Map lines delineate study areas and do not necessarily depict accepted national boundaries.

Two sensitivity analyses quantify the magnitude and direction of European calibration dominance. An equal-region-weighted Random Forest—with training weights inversely proportional to region label count—produced atlas predictions correlated at r = 0.91 with the primary model (MAD = 2.14 pp) and a headline PIF-based estimate of 10.13 million DALYs (+6.3%), indicating that regional training imbalance induces a modest conservative bias. A model trained exclusively on non-European labels (n = 21) predicted the 23 held-out European labels at R^2^ = 0.23 (MAE = 6.89 pp), produced a global atlas differing from the primary model by MAD = 2.75 pp (r = 0.89), and yielded a PIF-based estimate of 10.62 million DALYs (+11.4%), quantifying the directional impact of European calibration dominance. Both analyses indicate that the primary model is level-biased downward but structurally consistent; the bias lies within the leave-one-out residual interval and is addressed by the R^2^-weighted reliability estimate.

### Ecological associations between UPF supply and NCD outcomes

In crude between-country OLS, UPF supply share was associated with 22 of 27 outcomes at FDR < 0.05; 7 remained significant after GDP adjustment (Figure 2). Within-between (Mundlak) triangulation revealed marked between-country coefficients (6/9 outcomes, FDR < 0.05) but near-zero within-country effects (BMI: between +1.35 SD vs. within −0.05 SD; Hausman p < 0.05 for 6/9), consistent with the generational lag hypothesis. Spatial autocorrelation results are in Supplementary Table S5.

**Figure 2.** Ecological associations between UPF supply share and 27 NCD outcomes. Left panel: crude between-country OLS. Right panel: GDP-adjusted. Bars indicate per-1-SD standardised beta coefficients.

To assess residual spatial autocorrelation, we implemented Conley spatial HAC standard errors[44] at 2,000 and 4,000 km for three core outcomes. All three remained significant under spatial correction (p < 0.005 at both thresholds). Core associations for type 2 diabetes, cardiovascular disease, and BMI remained significant under Conley spatial HAC standard errors at both thresholds, indicating that observed spatial clustering reflects genuine regional dietary patterns rather than residual autocorrelation (Supplementary Table S5).

> *Most conservative (Layer I): 1.59M DALYs (T2D RR 1.06, CVD RR CI low 1.002, 25% waste, LAC excluded; does not include individual calibration). Fully conservative including individual calibration (Layer P, kappa=0.02): 0.19M. Individual calibration (O-P): k derived from 44x BMI ecological inflation converted to PIF scale; kappa=0.04 is the primary intake-adjusted estimate, kappa=0.02 the conservative lower bound. Layers C-F are exposure-side adjustments applied to the baseline (A); Layers G-H are RR-side adjustments; Layers I-Q are combined or cross-cutting scenarios. Layer I combines exposure-side and RR-side adjustments without individual calibration; Layer P adds kappa=0.02 to the Layer I scenario*.

### Potentially avoidable NCD burden in 2021

At TMREL = 15%, the supply-side PIF-based estimate yields 8.08 million DALYs (T2D 5.35M, CVD 2.73M) across 133 countries, excluding LAC where atlas generalisability failed (leave-region-out R^2^ = 0.00). Including LAC yields 9.53 million DALYs as an upper-bound sensitivity scenario (Table 2, Figure 3). Global-mean PIF: T2D 9.9% (95% UI 9.1–11.4%). The five-layer sensitivity cascade spans 1.59M (most conservative) to 27.97M (TMREL = 0%), with a credibility-weighted estimate of 3.62M. Income-stratified estimates appear in Supplementary Table S6.

**Figure 3.** PIF-based burden estimation. (A) Country-level T2D PIF, top and bottom 15 countries. (B) Four-component Oaxaca decomposition of burden growth, 2010-2021, showing denominator-driven and exposure-driven contributions. (C) TMREL sensitivity cascade across six threshold scenarios. (D) Overall sensitivity range from fully conservative (0.19M, including individual calibration) to theoretical upper bound (27.97M DALYs). The most conservative combined scenario without individual calibration (Layer I) is 1.59M.

The R^2^-weighted estimate of 3.62 million uses a floor weight of 0.01 for LAC countries (where leave-region-out R^2^ = 0.00). Sensitivity of this floor choice is modest: at floor = 0 (equivalent to LAC exclusion), the weighted estimate is 3.66 million; at floor = 0.05, 3.73 million; at floor = 0.10, 3.80 million; at floor = 0.20, 3.95 million (Supplementary Table S7). The primary estimate is thus robust to the choice of floor value across the plausible range [0, 0.20], varying by less than 10%.

At the primary TMREL of 15%, 26 of 166 countries (15.7%) fall below the threshold and have zero PIF for T2D—these countries account for 24.7% of total T2D DALYs and 33.4% of CVD DALYs in our panel, meaning that approximately one quarter of the global diabetes burden is structurally excluded from our PIF calculation by the TMREL cutoff alone. At TMREL = 10%, no countries fall below the threshold, and at TMREL = 20%, 57 countries (34.3%) are excluded, zeroing 54.2% of T2D DALYs. The theoretical upper bound at TMREL = 0% yields 27.97 million DALYs. These cutoff impacts highlight the structural sensitivity of PIF-based burden estimates to TMREL specification (full sensitivity in Supplementary Materials).

### Drivers of burden growth, 2010-2021

PIF-based burden rose from 5.98M (2010) to 8.08M (2021, LAC excluded), a 35% increase. Including LAC, the corresponding figures are 7.17M to 9.53M. Oaxaca decomposition attributes 103% to denominator expansion (population growth, ageing, epidemiological transition) and −3% to UPF supply change. Global-mean atlas UPF supply shifted +0.51 pp. Results are robust to sensitivity adjustments (Supplementary Table S7).

The observed global-mean change (+0.51 pp) is an order of magnitude smaller than the leave-one-out MAE (6.47 pp). Under a simple error-propagation framework, if prediction errors are independent and approximately normal, a 95% confidence interval for the true global-mean change would span approximately ±1.2 pp (standard error of the mean across 166 countries, incorporating imputation uncertainty). The Oaxaca exposure-driven component (−3%) is therefore statistically indistinguishable from zero and may plausibly range from −15% to +10% depending on error correlation structures. The near-zero exposure contribution is consistent with the structural stability of global UPF supply during 2010–2021 (Figure 4). Alternative decomposition sequences confirm this pattern (Supplementary Table S7-extended).

**Figure 4.** Regional UPF supply transition heterogeneity, 2010-2021. (A) Dumbbell plot of regional UPF supply share change by World Bank region. Filled circles denote 2021 values; hollow circles denote 2010 baselines. Colours indicate transition status: accelerating (red), slow growth (orange), high-level steady state (blue), and declining (green). (B) Net regional UPF supply change relative to the global mean (+0.51 pp, vertical dashed line). Only North America and Europe show increases exceeding 1 pp. All other regions cluster near zero or show slight declines, consistent with a structurally established global processed-food environment. Transition status defined by net change: accelerating (>1 pp), slow growth (0.5-1 pp), stable (<|0.5| pp), and declining (<-0.5 pp). South Asia (−0.9 pp) and Middle East & North Africa (−1.1 pp) are classified as declining rather than stable.

Regional heterogeneity beneath the global mean. The global-mean UPF supply change of +0.51 pp masks substantial variation (Figure 4). North America and Europe showed the largest increases (+4.7 pp and +2.6 pp), while most other regions showed near-zero or slight declines (East Asia & Pacific: −0.3 pp; South Asia: −0.9 pp; Middle East & North Africa: −1.1 pp). Regions showing acceleration already have the highest absolute UPF supply levels, while low-baseline regions (sub-Saharan Africa: 15.8%; South Asia: 19.4%) show stable or declining shares—implying differentiated policy architectures (discussed below as the “high-level equilibrium paradox”).

### Generational lag evidence from 60-year historical data

Processed macro-ingredient supply rose monotonically across all regions from 1961 to 2023 (Figure 5). The 1970s share correlated with 2022 obesity at r = 0.52 (n = 123, p < 0.001). Multi-lag fixed-effects results peak at ∼20 years: β = +0.024 (p = 0.024), consistent with a generational lag between dietary exposure and NCD manifestation.[15,43,45]

**Figure 5.** Generation-lag analysis. (A) Sixty-year processed macro-ingredient supply share by region, 1961-2023. (B) 1970s supply share versus 2022 adult obesity prevalence (r = 0.52, n = 123). (C) Multi-lag panel fixed-effects coefficients: obesity regressed on processed supply share at 10, 15, 20, and 25-year lags, showing peak effect at 20 years.

## Discussion

### Principal findings

We report a globally representative, time-varying assessment of UPF supply and potentially avoidable NCD burden with fully decomposed uncertainty. The atlas validates well in East Asia & Pacific (R^2^ = 0.94) and moderately in sub-Saharan Africa and Europe, but fails to generalise to LAC (R^2^ = 0.00). Given this generalisation failure, we exclude LAC from the policy-relevant baseline, yielding PIF-based estimates of 8.08M DALYs (range 1.59M most conservative to 8.08M baseline), with LAC-inclusive figures (9.53M) reported as an upper-bound sensitivity scenario and a credibility-weighted estimate of 3.62M.

### Interpretation and comparison with existing evidence

Our atlas provides the first open, version-controlled UPF supply dataset with quantified regional validity. Our exclusive reliance on supply-side features—excluding GDP and urbanisation from imputation—was a deliberate design choice: national UPF supply share operates as an aggregate indicator of food-system commercialisation, consistent with the CDoH view that transnational corporations shape diets through supply-chain reconfiguration rather than individual choice architectures alone.[24]

What supply share measures and omits in the CDoH framework is consequential. National UPF supply share captures the aggregate outcome of transnational food-system restructuring—commodity chain penetration, retail format transition, and ingredient trade flows—without disaggregating specific corporate actors. It is therefore a population-level exposure indicator consistent with CDoH theory, not a firm-level mechanism. The exclusion of GDP and urbanisation from the imputation model ensures that the atlas quantifies the commercial restructuring of food availability rather than general economic development. Future research should nest supply-share trends within firm-level panel data to distinguish transnational from domestic capital drivers.

Our T2D PIF of 5.6% of incident DALYs is roughly half the aggregate dietary risk PAF reported by O’Hearn et al. (2023), consistent with differences in exposure definition (supply vs. intake), outcome metric (DALYs vs. incident cases), TMREL specification (15% vs. 0%), and cross-level calibration (Supplementary Table S9).

> *The normative politics of TMREL. Our primary TMREL of 15% is calibrated to the lowest quartile of observed national intake surveys, which are predominantly European. This calibration is a pragmatic choice given available data, not a claim that 15% represents a universal biological threshold. For populations in the Global South, where traditional food systems and informal markets dominate, a 15% threshold derived from high-income dietary patterns may misclassify culturally integral processed foods as harmful exposure*. *We therefore treat TMREL as a policy-relevant feasible minimum rather than an ontological boundary, and report sensitivity across 10–20% to partially bound this geographic uncertainty. Future region-specific TMREL calibration should draw on local dietary survey data and culturally adapted food classification systems*.
>
> *Our PIF-based approach complements the GBD comparative risk assessment framework. GBD 2021 attributes approximately 188 million DALYs to suboptimal diet across 15 risk factors; our UPF-specific layer provides a counterfactual scenario currently absent from that hierarchy*.

A further conceptual limitation of the TMREL framework arises in the context of the double burden of malnutrition. In low- and middle-income countries where undernutrition and micronutrient deficiency coexist with rising NCD prevalence, ultra-processed foods may displace not only whole grains and fresh produce but also nutritionally inadequate traditional diets. Under these conditions, the counterfactual of reducing UPF supply to 15% of energy carries a different nutritional risk–benefit calculus than in high-income settings, where UPF predominantly displaces already adequate diets. The TMREL construct, derived from European intake distributions, does not internalise this trade-off. Future burden assessments should incorporate double-duty action frameworks [53] that simultaneously account for undernutrition and NCD risk, rather than treating UPF reduction as an unambiguous universal target.

### Public health and policy implications

Two policy-relevant estimands. We distinguish two estimands with distinct policy functions. The ecological PIF (8.08M DALYs, LAC excluded) serves as a structural upper bound for cross-country resource allocation and comparative priority-setting; it captures the maximum burden under the assumption that country-level supply changes translate one-for-one into population health, and excludes regions where the atlas cannot generalise. The individually calibrated estimate (0.19–0.48M DALYs) provides a plausible lower bound for individual risk communication and clinical expectation-setting. Policy discussions that ignore this approximately 20-fold interval risk overcommitting to exposure reduction in contexts where the true attributable burden is uncertain. We use the LAC-excluded ecological estimate as the primary reference for the remainder of the Discussion, while noting the full range in all quantitative claims.

The finding that recent burden growth is denominator-driven has direct implications for policy framing. It suggests a shift from emergency response to structural management is warranted

### Epistemic injustice and the geography of data

The atlas exposes a structural asymmetry in global nutrition knowledge: 55% of training labels come from Europe and Central Asia, while South Asia, the Middle East and North Africa, and Central Asia are essentially unrepresented. The LAC generalisation failure (R^2^ = 0.00) is instructive—the dominant predictor (sugar supply) is substantially weaker in LAC than elsewhere, reflecting dietary traditions where sugar is a less reliable UPF proxy. A Northern-derived measurement vocabulary is thus partially deaf to Southern dietary structures. We advance a data gap roadmap (Supplementary Table S2) for prioritising new NOVA survey collection in underrepresented regions.[23]

### Governance beyond the nation-state and beyond the electoral cycle

Our generational lag results reframe the temporal politics of UPF policy. The ∼20-year peak effect of processed-food supply on obesity, combined with near-zero exposure-driven burden growth, suggests—pending causal confirmation—that contemporary NCD burden partly reflects dietary environments established ∼20 years ago. Political cycles span 3–5 years; food-borne chronic disease cycles span ∼20 years. This structural mismatch means interventions evaluated on electoral timescales will routinely appear to fail.[15,18] The appropriate institutional response would be independent food-governance institutions with mandates longer than electoral terms and monitoring infrastructure tracking the intervention-to-outcome pipeline over decades.[17,20] Because UPF supply chains are transnational, governance also requires coordinating instruments beyond the nation-state. The atlas is a modest contribution to this longer-horizon institutional capacity. These governance implications are illustrative inferences drawn from the temporal structure of our epidemiological estimates; they are not derived from institutional or political feasibility analysis, which lies beyond the scope of this study.

### The high-level equilibrium paradox: differentiated policy architectures for a stratified world

The high-level equilibrium paradox: a hypothesis-generating framework for differentiated policy architectures. The regional heterogeneity in Figure 4 suggests a tentative pattern that we label the high-level equilibrium paradox—a hypothesis-generating interpretive framework rather than an established empirical law. In high-income countries, UPF supply has been structurally entrenched since before 2010, the population is ageing, and burden growth is denominator-driven. If this pattern is confirmed by future firm-level and retail-transition research, high-income countries may need to complement exposure-reduction policies with population-resilience strategies, whereas transitioning economies face a preventive-containment window. This interpretation draws on qualitative food-systems research documenting product-level upgrading and market-saturation strategies by transnational food corporations in high-income settings (Baker et al., 2020)[24], though we lack direct firm-level data to confirm these mechanisms. A single global narrative obscures the fact that appropriate policy architecture depends on where a country sits on the commercialisation trajectory. National means further mask within-country stratification: in high-UPF settings, denominator-driven burden growth may concentrate among ageing middle-class cohorts, while low-income groups continue to experience exposure-driven increases. Future multilevel designs should link national trade policy to neighbourhood retail infrastructure and individual consumption. We emphasise that this framework remains speculative pending direct evidence on corporate product portfolios, retail-format transitions, and within-country dietary stratification.

> *Two structural factors may explain the observed regional divergence. In high-UPF settings, incremental growth may reflect product-level upgrading by transnational corporations rather than increased market penetration. In low-UPF settings, stability may reflect barriers to commercial penetration (informal food sectors, tariff protection, limited cold-chain logistics). Regions with the largest tariff reductions over 2010–2021 saw UPF supply share increases approximately 2–3 percentage points higher than protected regions (Supplementary Figure S5)*.

## Limitations

This study has important constraints that condition interpretation. For each limitation, we describe its nature, the measures taken to address it, and the residual impact on our estimates.

> *Contextual omission. National mean UPF supply shares conceal subnational polarisation (higher in urban areas, younger cohorts, lower-income neighbourhoods). Because our outcomes are national aggregates, we cannot assess within-country distributional inequity, which we flag as a priority for future multilevel analysis*.
>
> *Epistemic coloniality. The NOVA classification was developed from and validated against dietary structures prevalent in high-income settings; applying it uniformly to 174 countries imposes a Northern-derived measurement framework that may misclassify culturally integral traditional products. The ML imputation compounds this by learning from a geographically skewed label set (55% European). Traditional fermented, dried, or culturally processed foods may be misclassified in non-Western contexts. Our CDoH and epistemic injustice frameworks serve as interpretive lenses; claims about mechanisms should be understood as theoretically grounded interpretation rather than causal identification*.
>
> *Ecological study design. Our between-country associations are subject to the ecological fallacy. We have directly quantified this inflation: the cross-level comparison shows a 44-fold discrepancy between ecological and individual-level BMI effects (Supplementary Materials). We report ecological associations as population-level patterns, not individual-level causal effects*.
>
> *Exposure measurement. UPF supply shares differ from individual dietary intake. We address this through waste-deduction sensitivity (producing a threefold range), intake calibration (within 5% of supply-side baseline), and consistent terminology. The processed macro-ingredient proxy (r = 0.66 with NOVA-UPF) captures the broader processed-food transition, not UPF-specific effects. The generational lag analysis uses adult obesity rather than T2D or CVD because historical NOVA-UPF data do not exist prior to 2003; we cannot exclude the possibility that T2D and CVD follow different latency distributions*.

The processed macro-ingredient proxy correlates with atlas NOVA-UPF at r = 0.66, implying that 44% of the variance in true UPF exposure is unmeasured in the historical panel. In fixed-effects regressions, such classical measurement error typically produces attenuation bias toward the null. The observed 20-year lag coefficient (β = +0.024, p = 0.024) may therefore underestimate the true generational lag effect, and the peak at ∼20 years should be interpreted as a conservative lower bound rather than an unbiased point estimate.

> *Regional atlas validity. Leave-region-out R-squared ranges from 0.00 (LAC) to 0.94 (EAP). We propagate regional reliability weights into all PIF estimates, report results with and without LAC exclusion, and explicitly identify priority regions for future NOVA survey collection—South Asia (1 label for 8 countries), the Middle East and North Africa (0 labels), sub-Saharan Africa beyond the current 4 labels, Central Asia (0 labels), and LAC. Additional methodological and theoretical references informing our interpretation are provided in the full reference list.[46–50]*
>
> *Data gap roadmap. Our atlas identifies five priority regions where additional national NOVA-UPF intake surveys would most improve global UPF monitoring: (i) South Asia (1 label for 8 countries, representing 1.9 billion people), (ii) the Middle East and North Africa (0 labels for 21 countries), (iii) sub-Saharan Africa beyond the current 4 labels (44 countries with 0 labels), (iv) Central Asia (0 labels), and (v) Latin America—not for label quantity (7 labels exist) but because the current FAO supply features fail to capture LAC UPF patterns (leave-region-out R^2^ = 0.00), suggesting that LAC-specific predictor variables (e.g., food import dependency, supermarket penetration, trade agreement indicators) may be needed*.
>
> *The LAC generalisation failure has a specific mechanism that quantitative diagnosis identifies: LAC countries have systematically higher sugar supply shares (27% vs 18% of kcal in non-LAC, p < 0.001), and the correlation between sugar share—the atlas’s dominant feature—and NOVA-UPF is substantially weaker in LAC (r = 0.31) than in non-LAC countries (r = 0.64), reflecting distinct dietary traditions (e.g., high beef consumption in Argentina, Brazil, and Uruguay) where sugar supply is a less reliable UPF proxy. The LAC generalisation failure is not merely a technical limitation of the sugar predictor; it exemplifies how a Northern-derived measurement vocabulary—in which sugar is a reliable marker of ultra-processing—becomes epistemically deaf to Southern dietary structures where sugar carries different culinary and nutritional meanings. This is not a defect to be corrected by better feature engineering alone, but a structural limit of applying globally uniform classification systems to culturally heterogeneous food systems. It constitutes the empirical core of the epistemic injustice concern that our theoretical framework identifies: the geographic gaps the atlas exposes are themselves evidence about whose food systems become legible to global monitoring and whose remain invisible to the dominant measurement apparatus. The open-source release of our atlas code, feature pipeline, and label database is designed to facilitate rapid ingestion of new labels as they are published, enabling iterative improvement of the global UPF map*.
>
> *Dose-response transportability. RRs originate predominantly from high-income-country cohorts.[51–54] We address this through the 3×3 joint RR sensitivity grid (spanning 3.78-16.94 million), TMREL sensitivity (10-20%), and explicit acknowledgment that no LMIC-specific UPF dose-response estimates have been published. The Monte Carlo uncertainty intervals incorporate RR confidence interval variation. This is complemented by mechanism and intake-trend studies,[55–56] the policy and syndemic literature,[17,20] and cohort evidence on UPF intake and anthropometric, cancer, and mortality outcomes.[57–60]*
>
> *Effect modification by development context. The dose-response relative risks from Lane et al. (2024) are derived predominantly from high-income-country cohorts, where UPF typically substitutes whole grains, fresh produce, and home-prepared meals. In low- and middle-income settings, UPF may substitute nutritionally inadequate traditional diets, with potentially different net metabolic consequences. The direction of this effect-modification bias is theoretically ambiguous: in populations undergoing early nutrition transition, UPF might improve acute nutritional status while simultaneously increasing long-term NCD risk, yielding attenuated dose-response slopes relative to HIC settings; conversely, populations with higher baseline metabolic vulnerability due to foetal and early-life undernutrition may exhibit steeper dose-response gradients. Income-stratified estimates (Supplementary Table S6-extended) confirm that high-income countries bear the largest absolute PIF-based DALY burden, while low- and middle-income-country estimates carry the greatest uncertainty due to both weaker atlas validity in several regions and unverified RR transportability. Until LMIC-specific UPF dose-response data become available, our RR sensitivity grid (3×3 joint T2DxCVD, Table 2) partially bounds this uncertainty, and the individually calibrated estimates (kappa=0.02-0.04) further contract the plausible range. We encourage prospective cohort studies in LMIC settings to resolve this critical evidence gap*.
>
> *Training sample composition. European labels constitute 51% of the training set, a reflection of the geographic distribution of published NOVA surveys rather than a design choice. We disclose this imbalance, quantify its directional impact through training-set sensitivity analyses (Supplementary Table S4), and use leave-region-out cross-validation to assess generalisability. We explicitly disclose these constraints to define the evidential boundary of our estimates, rather than to invalidate the overall findings*.
>
> *Cross-level calibration. The individual-level calibration factor (k=0.04) is derived from US NHANES data under the assumption that the cross-level bias structure is transportable. This is a strong assumption: countries with lower UPF variance may exhibit smaller ecological inflation. No nationally representative individual-level NOVA data exist for 142 of 166 countries in our panel. The k-calibrated estimate (0.19-0.48M) should be interpreted as a sensitivity bound rather than a definitive individual-level burden estimate; the true burden likely lies between the individually calibrated lower bound and the ecological upper bound (9.53M)*.

The direction and magnitude of the ecological inflation captured by kappa likely varies by national development context. In high-income settings, UPF displaces whole grains, fresh produce, and home-prepared meals, producing a steep dose-response gradient. In low- and middle-income countries undergoing nutrition transition, UPF may partially substitute nutritionally inadequate traditional diets alongside nutrient-dense foods, yielding a more attenuated net metabolic effect. This implies that the 44x BMI ecological inflation factor observed in NHANES, and the derived kappa=0.04, may represent a lower-bound compression factor for LMIC populations, where the ecological-to-individual gap may be narrower and the kappa would be correspondingly larger. Until multi-country individual-level NOVA data become available, the kappa-calibrated estimates (0.19-0.48M) should be interpreted as a sensitivity scenario conditional on US dietary substitution patterns.

> *Dietary substitution and competing risks. Our counterfactual scenario assumes that reduced UPF supply is not offset by increased consumption of other unhealthy food groups (e.g., processed meat, refined grains). We do not model dietary substitution effects because nationally representative time-series data on complete dietary patterns are unavailable for most countries. Future burden assessments should incorporate multi-risk substitution frameworks as data permit*.

## Conclusions

We present a cross-validated global atlas of UPF supply and a time-varying PIF-based estimation of potentially avoidable NCD burden, with fully transparent uncertainty decomposition. The credibility-weighted estimate of 3.6 million DALYs is best interpreted as a mid-range scenario within a broad uncertainty interval spanning 0.19 million (fully conservative, individually calibrated) to 27.97 million (theoretical upper bound under TMREL = 0%). The most policy-relevant lower bound, combining adverse assumptions without individual calibration, is 1.59 million, and the supply-side baseline is 8.08 million (LAC excluded, reflecting zero regional generalisability), with LAC-inclusive sensitivity at 9.53 million. These figures represent counterfactual model scenarios, not directly realisable policy returns, particularly given the approximately 20-year generational lag between exposure change and health outcome manifestation. The processed-food transition was structurally established by 2010 in most countries, and contemporary NCD burden overwhelmingly reflects past rather than current dietary exposures. Policy evaluation should therefore adopt generational rather than electoral time horizons. The open atlas, sensitivity framework, and analytical code are released as public-health monitoring infrastructure.

## Data Availability Statement

All analytical code, processed UPF atlas datasets, and full sensitivity analysis outputs generated in this study are permanently archived on Zenodo[61] with a persistent DOI: 10.5281/zenodo.21785121. The code repository is also maintained on GitHub for community reuse: https://github.com/ShengZe-123/upf-atlas-global.

## Ethics Approval and Consent to Participate

This study used exclusively publicly available, aggregated and de-identified data (FAO Food Balance Sheets, GBD estimates, NOVA label datasets, and open food label repositories) and did not involve human participants, their private data, or biological samples. Ethical approval and informed consent were therefore not required, consistent with institutional and journal policy. Consent for publication is not applicable.

## Supplementary material

Supplementary Tables S1-S11 (including S3-extended for model benchmarking, S6-extended for income-stratified PIF reporting, and S7-extended for Oaxaca decomposition sequence sensitivity), Figures S1-S5, and the full analysis code repository accompany this manuscript. Contents include: complete 27-outcome Eco-PheWAS results (S1); atlas validation and ML benchmark results (S2-S4); spatial autocorrelation diagnostics including Conley SE (S5); income-group-stratified burden estimates (S6); Oaxaca decomposition sensitivity (S7); SSB tax synthetic-control per-country results (S8); Wang et al. 2022 decomposition and cross-level calibration tables (S9); and generational lag full panel regression diagnostics (S10). All materials are released under open-access terms.

## Author Contributions

S.H. designed the study, acquired public data, built statistical and machine learning models, conducted formal data analysis, and drafted the manuscript. X.W. contributed to data curation, validation, and manuscript revision. All authors approved the final manuscript and take full accountability for the content.

## Acknowledgements

No funding was received. We thank the FAO, WHO, GBD collaborators, and Open Food Facts for making their data publicly available.

